# Host genotypes interact with microbial communities to modulate gene expression in the human intestine

**DOI:** 10.64898/2026.01.30.26344673

**Authors:** Shreya Nirmalan, Sabrina Arif, Julong Wei, Sambhawa Priya, Ran Blekhman, Roger Pique-Regi, Francesca Luca

**Affiliations:** Center for Molecular Medicine and Genetics, Wayne State University, Detroit, MI; Section of Genetic Medicine, Department of Medicine, University of Chicago, Chicago, IL; Department of Obstetrics and Gynecology, Wayne State University, Detroit, MI; Department of Human Genetics, University of Chicago, Chicago, IL

## Abstract

**Background:** Inflammatory Bowel Disease (IBD) is characterized by chronic intestinal inflammation and is associated with both altered gut microbiome composition and host genetic risk. Both host genetic variants and the gut microbiome can affect host gene expression in the colon; however, it remains unclear whether interactions between the two (genotype x microbiome, GxM) shape intestinal gene regulation in humans and their contribution to IBD risk.

**Methods:** We analyzed publicly available data for 86 individuals (64 patients with IBD and 22 controls) in the Inflammatory Bowel Disease Multi’omics Database consisting of host genotype, host gene expression, and mucosal gut microbiome (16S rRNA) data from rectal and ileum biopsies. We performed expression Quantitative Trait Locus (eQTL) mapping and then used computational fine-mapping to identify likely causal variants. We tested whether microbial taxa modify genetic effects on host gene expression. We then integrated GxM eQTLs with IBD, Crohn’s Disease (CD) and Ulcerative Colitis (UC) Genome-Wide Association Study results by leveraging Transcriptome-Wide Association Studies and colocalization methods.

**Results:** We found 3,777 and 3,694 host genes with eQTLs in the rectum and in the ileum, respectively (FDR = 10%). Using the fine-mapped eQTLs, we found 36 GxM interactions for 31 host genes with 22 microbial taxa in the rectum and 30 GxM interactions in the ileum for 15 host genes and 20 taxa (FDR = 10%). Taxa with GxM interactions clustered into two distinct groups with opposing effects on host gene regulation and reflected distinct functions of microbes in the gut. i.e, butyrate producers versus sulfate reducers. Integration with IBD GWAS revealed that 23 variants with GxM regulated the expression of host genes putatively causal for IBD, CD or UC (FDR = 10%), thus identifying microbes that can either amplify or buffer genetic risk.

**Conclusions:** Our results show evidence of genetic effects on host gene expression that are modulated by microbiome composition, and provide insight into how IBD risk could be reduced by targeting specific microbial taxa contingent on host genotype.

## Background

Inflammatory Bowel Disease (IBD) is a group of disorders characterized by chronic inflammation and scarring of either the entire digestive tract (Crohn’s Disease, CD) or specifically the colon (Ulcerative Colitis, UC)[1,2]. IBD occurs due to an interplay between host genetic and environmental factors[3]. It is characterized by dysfunction of the intestinal epithelial lining and abnormal immune response to intestinal microbiota[4]. Current standard of care for IBD involves costly treatments including immunosuppressants and opioids, with many individuals still facing prominent disability and morbidity[5,6]. IBD has a substantial host genetic component, with over 200 genetic loci associated with IBD risk[7]. Some of these variants are in protein coding genes that play a role in host immune system engagement with bacteria and potential modulation of the microbiome[8–11]. However, the majority of IBD risk loci are in non-coding regions, suggesting that similar to other complex traits, alterations in gene regulatory processes are key mechanisms underlying IBD pathogenesis[12]. Indeed, several studies have shown that single nucleotide polymorphisms (SNPs) associated with interindividual variation in host gene expression (i.e. expression Quantitative Trait Loci or eQTLs) overlap with variants associated with complex traits in GWAS[13–16]. Mapping eQTLs to disease variants identified in GWAS therefore provides mechanistic insights into risk and progression of complex diseases including IBD[13,14,17].

The gut microbiota, which is the community of microbes residing in the gut, has critical impacts on the immunological, nutritional and energetic landscape of the host[18,19]. In addition to host genetic variation, shifts in gut microbiome composition are also associated with variation in host gene expression both *in vivo* and in *in vitro* human cell models[20,21]. Studies in mouse models have similarly demonstrated that microbiome composition can drive host transcriptional responses[22,23]. For example, colonization of segmented filamentous bacteria induces Th17 cell differentiation and enhances resistance to bacterial pathogens in germ-free mice[24]. Individuals with IBD have dysbiotic microbiome profiles characterized by increased levels of facultative anaerobes such as *E.coli* and decreased levels of obligate anaerobes including *Faecalibacterium prausnitzii[25]*. Dysbiosis in IBD is accompanied by dysregulation of several microbial processes, including carbohydrate metabolism, short chain fatty acid synthesis, and virulence[25–27]. Previous studies have also demonstrated that differentially abundant microbial taxa in IBD correlate with expression of host genes associated with the disease[25,28].

While progress has been made in understanding the host genetic and microbial factors associated with IBD, we are still missing an understanding of how microbiome composition shifts interact with host genetics to impact host gene expression and ultimately lead to disease. Genetic variants that influence transcriptional response to environmental changes (including chemicals, drugs and microbial agents[29–39]) represent molecular genotype-environment interactions (GxE) [29–35]. Several studies have demonstrated that these molecular GxE contribute significantly to variation in complex traits[30–32,35,40,41]. When we consider the intestinal microenvironment, characterized by the presence of distinct microbial communities, previous studies have not considered interactions between these communities and host genetic variation and their effect on host gene expression. We hypothesized that host genetic variants could interact with microbiome composition (i.e. Genotype x Microbe interactions (GxM)) to influence host gene expression. Here, we performed interaction eQTL mapping using paired host genotype, host gene expression and microbial abundance data to identify GxM interactions in the ileum and rectum (**Fig 1B**). We integrated GxM with IBD GWAS and TWAS data to determine how these interactions modulate IBD risk providing a novel avenue to understand the pathophysiology of this disease as well as to discover new therapeutic targets.

**Figure 1.**
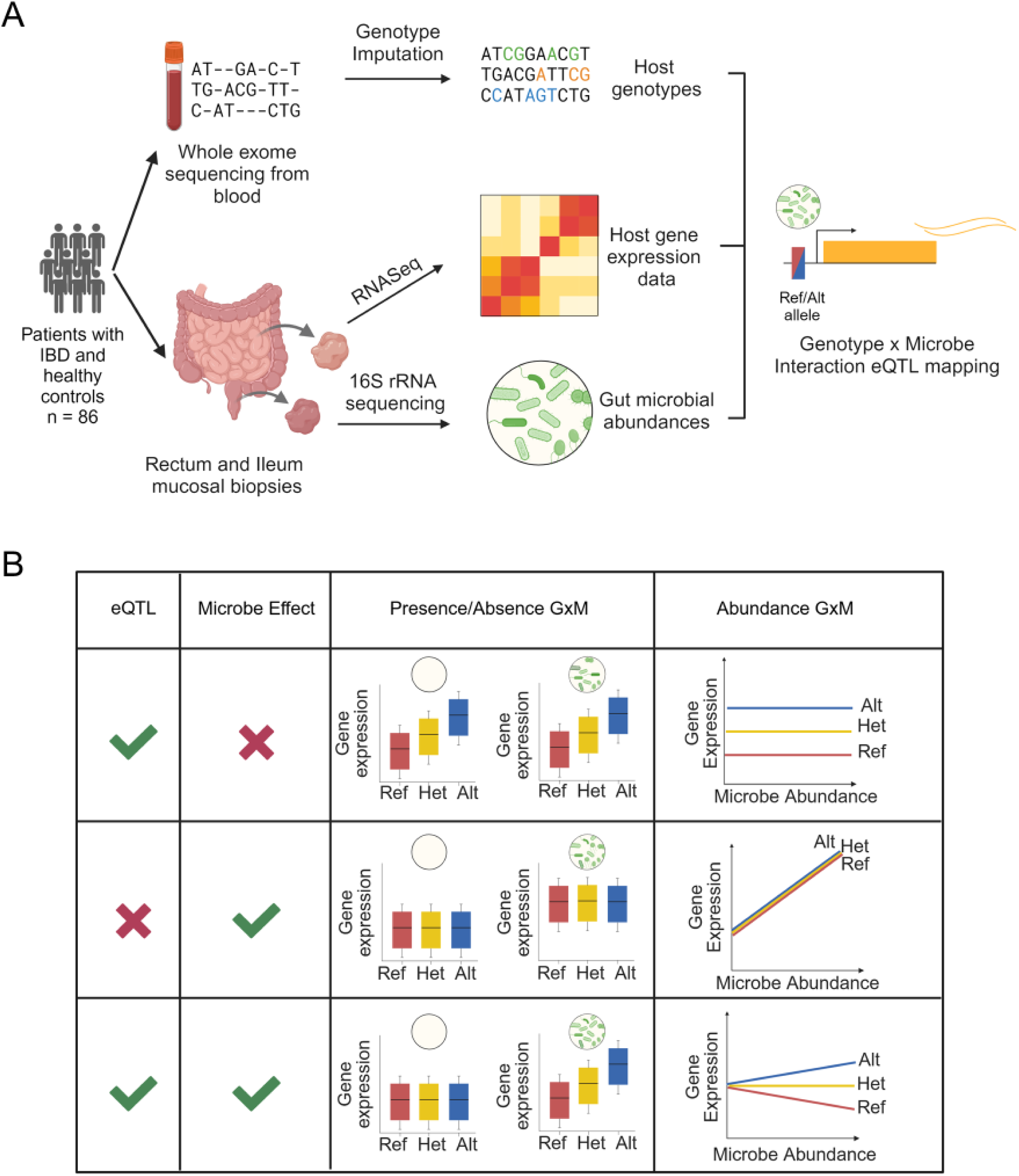
Study Design and Conceptual Framework. **A)** Study Design. **B)** Conceptual framework of how host genetic variation and microbiome composition can influence host gene regulation independently or through an interaction (GxM). The first two columns indicate if there is a eQTL (genetic effect) and/or a microbial effect on host gene expression. The third and fourth columns highlight how host gene expression could be impacted by host genotype and with the presence or absence of a microbe or microbial abundance depending on if there is a eQTL, microbial effect on host gene expression or an interaction.

## Methods

### Overall study design and data

For this study we used previously generated and published whole exome sequencing, and paired host gene expression (RNA-seq) and mucosal gut microbiome (16S rRNA) data from the IBD cohort that is part of the Human Microbiome Project (dbGaP Study Accession: phs001626.v1.p1). The original study obtained written informed consent from study participants and details of randomization and blinding during data collection is outlined in the publication of the original study[25,42].

The original cohort consisted of samples from 132 individuals. Samples were collected across 5 different sites with both pediatric and adult populations (ages 6 -76 years old). Detailed protocols for sample collection and processing are described at http://ibdmdb.org/protocols. For the rectum eQTL analyses, we focused on a set of 86 individuals (43 females, 43 males) with both whole exome sequencing and RNA-seq data collected from rectal biopsies. This cohort comprised 64 patients with IBD and 22 individuals without IBD (nonIBD). Out of the 64 patients with IBD, 40 patients had Crohn’s Disease, and 24 patients had Ulcerative Colitis. For interaction eQTL mapping, we obtained paired host RNA-seq and microbial abundance data derived from 16S rRNA-sequencing for 70 of the 86 rectum samples. For the ileum eQTL analyses, we used paired host RNA-seq and genotype data for 82 individuals (out of the 86 used for the rectum analyses). The ileum cohort consisted of 62 patients with IBD and 20 nonIBD individuals. Out of the 62 patients with IBD, 40 had Crohn’s Disease and 22 had Ulcerative Colitis. For interaction eQTL mapping in the ileum, we used paired host RNA-seq and microbial abundance data from biopsies for 69 of these individuals.

Detailed cohort characteristics for this dataset are included in Supplementary Table 1. Host gene expression data and whole exome sequencing data were accessed through dbGaP and downloaded as SRA files in December 2022. Then SRA files were converted to fastq files using sratoolkit[43]. We downloaded the processed OTU tables for the microbial data publicly available from the IBDMBD website (http://ibdmdb.org/).

### RNA Seq data processing

HISAT2[44] was used to align demultiplexed RNA-seq reads to the human genome version HG38 (‘GRCh38_snp_tran’) which considered splicing isoforms and common genetic variants. We then used samtools[45] to remove PCR duplicates and further removed reads with a quality score of <10 (equating to reads mapped to multiple locations). Aligned and cleaned (deduplicated) reads were counted using HTSeq[46] and the GRCh38 transcriptome assembly across 60,612 genes. Post-sequencing quality control included genotype QC check against respective DNA samples. All samples passed this QC step.

To finalize the selection of RNA samples used for this study we first selected individuals with transcriptomic data from the rectal biopsies (n individuals = 90, number of samples = 100). We then removed samples with low read counts (< 10^7^) or missing data for age or without corresponding genotype data, leaving 86 samples from 86 individuals for the rectum and 82 samples/individuals for the Ileum. For individuals with more than one rectum sample we used the sample with the highest fraction of reads (after removing duplicate reads) for our analyses. We filtered the gene expression data to include only protein coding genes and genes present on autosomes, and that were expressed with counts per million (CPM) > 0.1 in at least 20% of samples. The final processed RNA-seq dataset for the Rectum analyses contained 86 unique samples and 15,388 genes. For the Ileum analyses, we used the same approach to select the RNA-seq sample for individuals who had multiple ileum samples and used the same filtering steps, and thus the Ileum RNA-seq dataset contained 82 unique samples and 15,508 genes.

### Microbial data processing

The processed gut microbial data generated in Priya et al 2022[28] was used for this study for the rectum analyses. They subsetted the dataset to include data from colonic biopsy samples (rectum, sigmoid colon and transverse colon). For this study, we subsetted the dataset to the 70 rectum samples (out of 86) with 16S data for 121 taxonomy levels. For the Ileum analysis, we processed the microbial data generated from the 16S rRNA-seq data from the ileum biopsies for 72 of the 82 individuals using the same approach as described in Priya et al 2022. Briefly, we downloaded the unnormalized and unfiltered operational taxonomic unit (OTU) table of the 16S rRNA data. Sequences classified as Archaea, from chloroplasts, known laboratory-related or environmental contaminants were removed from the unnormalized OTU table. Next, the OTU table was summarized at the species (if present), genus, family, order, class and phylum taxonomic levels, and then was filtered to retain taxa found at 0.001 relative abundance in at least 10% of the samples. To allow for identification of associations at any taxonomic level without repeating analyses at each taxonomic rank, summarized taxa matrices at different ranks were combined into a taxa matrix resulting in a total of 115 taxa to be tested. The microbial data was then transformed using a centered log ratio (CLR) transformation across all 73 samples.

### Genotype data

Individuals were genotyped using whole exome sequencing data and imputed using the 1000 Genomes database[47] by Gencove (New York, NY). These data were used for sample quality control (see Genotype QC and Sex QC) and to perform eQTL mapping. The top three genotype principal components (PCs) were calculated to use as covariates in all statistical analyses.

### Genotype QC

To ensure that we correctly matched host gene expression and genotype data, we compared genotypes of RNA and DNA from all individuals used in this study. We used samtools[45] mpileup to call genotypes from RNA-seq BAM files for NCBI dbSNP[48] Build 144 variants. We used plink2[49] -make-king-table function to calculate a KING-robust kinship estimator to compare genotype calls across all DNA and RNA samples. All RNA samples passed this QC filter. Ultimately, the pairwise kinship coefficient between genotype calls from RNA and their respective DNA samples from the same individual ranged between -0.06 and 0.14. In contrast, the pairwise kinship coefficient rate between all other unrelated samples ranged from -1.59 to -0.39. This rate is not a direct measure of genotype call accuracy but is useful to identify possible sample swaps and mismatches.

### Sex QC

To check consistency of self-reported sex against genetic data, we plotted the fraction of reads in the RNA-seq data mapping to the Y chromosome for all samples. We noted a clear separation between the sexes with no outliers (**Supplemental Figure 3**).

### cis-eQTL mapping and enrichment in GTEx

We calculated gene expression residuals for both the rectum and ileum analyses by first performing CPM normalization to account for sequencing depth and then quantile normalizing the cpm normalized gene expression data across individuals for each gene using the qqnorm() function in R; second, we regressed out the following confounding variables: sex, age, percent reads mapping to exons, genotype PC1, genotype PC2 and genotype PC3 by fitting a linear model and then extracting the gene expression residuals. We then used FastQTL[50] with adaptive permutations (1000 - 10,000) for eQTL mapping. For each gene, we tested all genetic variants within 100Kb of the transcription start site (TSS) and with a cohort MAF > 0.1, for a total of 15,388 genes and 53,695,921 variant-gene pairs tested for the rectum and 15,508 genes and 54,285,257 variant-gene pairs tested for the ileum. We optimized the number of gene expression PCs included in the model to maximize the number of eGenes. The model that yielded the largest number of eGenes for the Rectum included 16 gene expression PCs and the model that yielded the largest number of eGenes for the Ileum included 17 gene expression PCs. Supplemental tables 2 and 4 contain the full eQTL analysis results for the lead SNP tested for each gene for the rectum and ileum.

We then used data from the GTEx cohort for the transverse colon (368 individuals), sigmoid colon (318 individuals) and terminal ileum (174 individuals)[51] (phs000424.v8) to check for enrichment of the IBD rectum and ileum eGenes in these datasets. The summary statistics of these datasets were obtained by running FastQTL software and testing genetic variants within 1 Mb of the TSS for each gene. Using the union of all eGenes tested in all 5 datasets we created an Upset plot using UpsetR[52] to visualize the overlap between them. Then we performed Fisher’s exact tests on 2 × 2 contingency tables using the set of eGenes tested between either the IBD rectum or IBD ileum eGenes and the three GTEx eGene datasets, indicating whether a gene harbored an eQTL (FDR = 10%) in the IBD Ileum or Rectum data (yes/no) and if the gene harbored an eQTL in tested GTEx dataset (FDR = 5%) (yes/no).

### Interaction eQTL mapping

To identify interaction eQTLs, we performed fine-mapping of the rectum and ileum eQTL signals using DAP-G, a Bayesian multi-SNP association model with a deterministic approximation of posteriors (DAP) algorithm[53] on a 1Mb window around each TSS. We then selected the SNP with the top PIP from fine-mapping as the lead eQTL to test for interaction with the microbiome for the eGenes found using FastQTL. Supplemental tables 3 and 4 contain the SNP-gene pairs used for the interaction model for the rectum and ileum. We considered two potential models of interaction based on the microbial abundance: the presence/absence of the microbe in the overall microbiome, or the relative abundance of the microbe.

In order to determine if a microbe was present or absent, we started from the untransformed OTU table and considered a microbe to be present if its relative abundance was > 0 for at least 25% of samples, indicating it could be present in at least one of the disease status subgroups (Non IBD, Crohn’s disease or Ulcerative Colitis). For the rectum microbial data, out of the 121 taxa analyzed, 45 taxa met these criteria and for the ileum, 65 taxa met these criteria.

For the presence/absence interaction model, we fit a linear model that includes both the genotype dosage and the marginal effect of the presence of a microbe (0 = absent, 1 = present), as well as their interaction to model the host gene expression residuals (see above): Expression ∼ genotype dosage + absence/presence microbe + genotype dosage:absence/presence microbe. To fit this model, we used the lm function in R-3.5.2. Storey’s q-value method to control for FDR was applied on the interaction p-values for all tests within each taxon separately. Supplemental Tables 6 and 7 contain the full results for the interaction eQTL presence/absence analysis for the rectum and ileum.

Interaction eQTL mapping for microbial abundances was performed in the same way as with the presence/absence model, except the model included the CLR-transformed abundance of the microbe: Expression ∼ genotype dosage + microbial abundance + genotype dosage:microbial abundance. For this model we tested for GxM with 121 taxa from the rectum 16S rRNA data and 115 taxa from the ileum 16S rRNA data. Supplemental Tables 8 and 9 contain the full results for the interaction eQTL presence/absence analysis for the rectum and ileum.

### Integration of eQTL, GWAS and GxM data for IBD risk

We used the fine-mapped eQTLs from the ileum and rectum data along with fine-mapped IBD GWAS data from TORUS as input to perform colocalization analysis using fastenloc[54]. We obtained gene locus-level colocalization probability (GLCP) for IBD risk. We then conducted a TWAS analysis to identify genetically regulated gene expression correlated with IBD risk by integration of the eQTL mapping results with IBD GWAS. We used the GWAS summary data for Inflammatory Bowel Disease, Crohn’s Disease and Ulcerative Colitis from the UK Biobank[55], including 8,873,545 genetic variants in autosomes. Similar to the SMR approach[56], we selected the top eQTL for each gene based on the highest PIP from the fine-mapping results obtained using DAP-G. In some cases, the top eQTL was not tested in the GWAS. In that case, we selected the closest SNP (within 100 Kb) in the GWAS. We then calculated the SMR p-value as previously reported[56] and used the q-value function to control for the false discovery rate to correct for multiple testing. IBD associated genes by TWAS are defined as those with FDR < 10%. Using the INTACT R package[57] we combined the colocalization results (GLCP) and TWAS z-score to derive the probability of putative causal genes for IBD risk. The putative causal genes are defined as those with FDR < 10% from INTACT. Supplemental tables 10 and 11 contain the significant IBD risk genes identified in the rectum and ileum. We then used a stratified FDR approach[58] to identify GxM harbored in IBD risk genes by re-calculating the Benjamini-Hochberg p-adjusted value for each of the identified genes in each body site for both the abundance GxM and presence/absence GxM results. Supplemental tables 12 and 13 contain the significant IBD risk genes with GxM results.

GWAS summary statistics report the effect of the alternate allele on disease risk where sign(GWAS) > 0 indicates that the alternate allele is associated with increased disease risk whereas sign(GWAS) < 0 indicates that the alternate allele is protective. Similarly, eQL effects from FastQTL are reported relative to the alternate allele, such that sign(eQTL) specifies whether the alternate allele increases or decreases gene expression. The TWAS direction for each gene indicates whether increased genetically predicted gene expression is associated with increased disease risk (positive sign) or decreased risk (negative sign). Under a single causal variant assumption (SMR), the TWAS direction was calculated as: sign(TWAS) = sign(GWAS) x sign(eQTL). To determine whether each microbial taxon increased or decreased risk in the context of its interaction with host genotype, we considered the effect of the alternate allele on risk from GWAS and the direction of the GxM interaction effect. For each gene-taxon pair, we computed a taxon-specific risk-direction score as: Taxon effect on risk = sign(GWAS) x sign(interaction effect), where sign(interaction effect) is the sign of the GxM interaction term (genotype dosage:microbial abundance) and represents whether increased microbial abundance amplifies or buffers the effect of the alternate allele on gene expression. A positive sign indicates that increased abundance of the taxa amplifies the effect of the risk allele (text is colored in red), whereas a negative sign indicates that the taxon buffers the effect of the risk allele (text is colored blue).

### Overlap with single-cell IBD dataset

We downloaded the seurat object for the full gene expression matrix of the scIBD dataset that is publicly available[59]. Seurat (V4)[60] was used for visualizing the scRNA-seq data. Briefly, the dataset consists of scRNA-seq data for 101 cell types across the 9 major cell compartments for individuals with IBD and healthy controls, meta-analyzed across 12 different studies. We used the Seurat:Dotplot function to get the normalized average expression values and percent expressed values of the genes with GxM across both the ileum and rectum and within the rectum eGenes and ileum eGenes separately, in each of these major cell types for plotting purposes and to perform the Fisher’s enrichment test (**Supplemental Figure 5**). To this end, we performed Fisher’s exact tests on 2 × 2 contingency tables using the set of eGenes indicating whether a gene had a GxM (yes/no) and was highly expressed in the major cell compartment, defined as having a normalized averaged expression > 2 and expressed in at least 10% of cells in one of the sub cell types within this group (yes/no).

## Results

### Genetic effects on host gene expression in the rectum and ileum

To identify host genetic variants that regulate the response to the microbiome (GxM) we first performed eQTL mapping in rectum and ileum tissues from IBD patients and healthy controls. To this end, we used genotypes derived from whole exome sequencing, gene expression from RNAseq and microbiome abundances derived from 16S rRNA sequencing data for 86 individuals that were collected as part of the Inflammatory Bowel Disease Multi’omics Database (IBDMDB)[25] effort of the Integrative Human Microbiome Project. We first used genotypes with host gene expression data from rectal and ileum biopsies to perform eQTL mapping (**Fig 1A Study Design**). We defined an eGene as a host gene with at least one *cis*-eQTL. We identified 3,777 eGenes in the rectum at a false discovery rate (FDR) of 10% (**supp figure 1A**) and 3,694 eGenes in the ileum (**supp figure 1B**). We found 2,038 host genes that harbored eQTLs in both body sites, but only 310 SNP-gene pairs in common between the two body sites. We did however find that effects of eQTLs are significantly correlated between the two body sites (**Fig 2A**, R = 0.678, p-value < 2.2 * 10^-16^). This suggests a similar genetic regulatory architecture in both the ileum and rectum.

**Figure 2.**
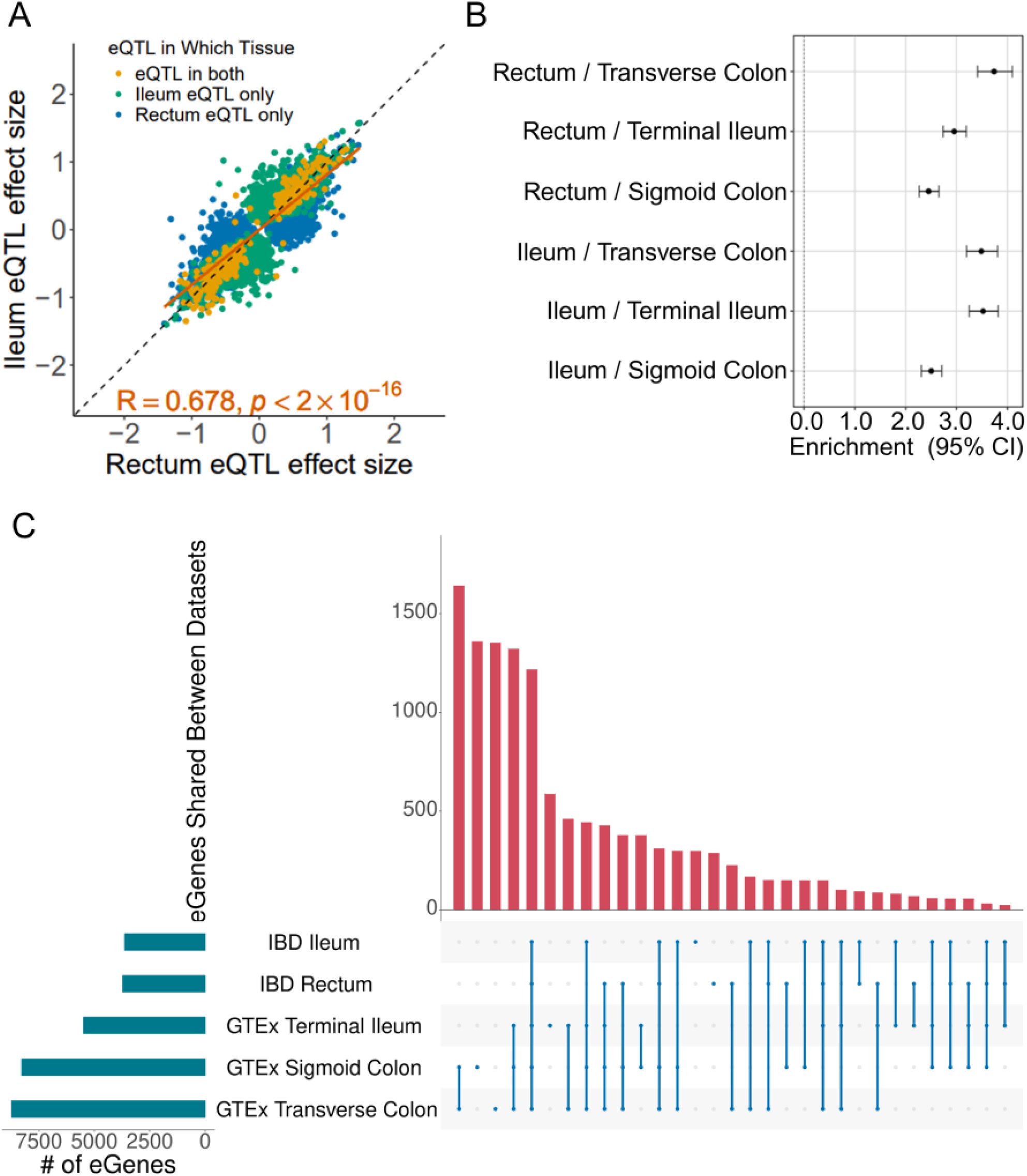
Rectum and ileum eQTLs are enriched in colon and ileum GTEx. **A)** Scatterplot of the effect sizes, of the union of gene-SNP pairs with significant eQTLs in either the Rectum or Ileum datasets. The Pearson correlation between these effect sizes is in red text at the bottom of the figure. **B)** Fisher’s exact test (p<2e-16) of enrichment of either the Rectum and Ileum eGenes in GTEx Transverse Colon, Terminal Ileum and Sigmoid Colon datasets. Error bars indicate 95% confidence intervals. **C)** Upset plot detailing overlap of eGenes between IBD Rectum and Ileum datasets with GTEx Transverse Colon, Sigmoid Colon and Terminal Ileum. The barplot on the left indicates the number of eGenes in identified in each dataset (FDR = 10%) and the barplot at the top of the figure indicates the number of eGenes shared between the datasets indicated by the combination matrix below the barplot.

In order to validate our findings, we queried if these eGenes were replicated in other larger eQTL cohorts, specifically the Sigmoid Colon, Transverse Colon, and Terminal Ileum datasets from the Genotype-Tissue Expression Project (GTEx)[61]. We found that the rectum eGenes were enriched in the GTEx sigmoid colon (Fisher’s test OR = 2.45 p-value < 2.2 * 10^-16^), transverse colon (Fisher’s test OR = 3.73, p-value < 2.2 * 10^-16^) and terminal ileum eGenes (Fisher’s test OR = 2.95, p-value < 2.2 * 10^-16^). Similarly, the ileum eGenes were enriched in the GTEx terminal ileum (Fisher’s test OR = 3.52, p-value < 2.2 * 10^-16^) sigmoid colon (Fisher’s test OR = 2.50, p-value < 2.2 * 10^-16^) and transverse colon eGenes (Fisher’s test OR = 3.49, p-value < 2.2 * 10^-16^) (**Fig 2B**). Of the eGenes identified, 1,218 were shared across all body sites, yet 298 eGenes were unique to the IBDMDB ileum dataset and 288 eGenes were unique to the IBDMDB rectum dataset (**Fig 2C**).

### Interactions between host genotype and gut microbial abundance highlight two clusters of taxa with opposite effects on host gene regulation

Having identified host genetic effects on host gene expression, we investigated whether these genetic effects vary with different microbial abundance across individuals. To this end we characterized host genotype x microbiome interactions (GxM), which we defined as cases where the relationship between microbial abundance and host gene expression varies with host genotype at a regulatory variant, or vice-versa, where genetic effects on gene expression vary with microbial abundance. To identify GxM, we first performed computational fine-mapping of the ileum and rectum eQTLs to identify the most likely causal variants for each eGene. Then we tested for interaction between host genotype at fine-mapped eQTLs and microbial abundance using a linear model. We found 24 interactions (GxM) for 23 host genes and 14 taxa in the rectum and 23 GxM for 23 host genes and 13 taxa in the ileum (FDR = 10%).

Prior work has identified groups of microbial taxa that act in a coordinated fashion on host gene expression[28]. We hypothesized that taxa may also exhibit coordinated patterns of GxM effects across host genes. To test this hypothesis, we calculated the correlation of the interaction effects between pairs of microbial taxa with GxM and we found two distinct clusters of taxa that have opposing interaction effects in each body site (**Fig 3A, D**). In the rectum, cluster A contained phylogenetically diverse taxa of mixed metabolic functions in the host; there were short chain fatty acid (SCFA) producers including *Butyricicoccus* which is found to be in lower abundance in individuals with IBD[62], and mucin degraders like *Sutterella* and *Bilophila* which both have pro-inflammatory capabilities[63,64]. Rectum-cluster B contained all taxa belonging to the phylum *Firmicutes* that are involved in fiber fermentation and SCFA producers and usually found to be in decreased abundance in individuals in IBD[65–69] (**Fig 3A**). *Butyricicoccus* in cluster A and *Dialister* in cluster B, while both are SCFA producers, interestingly had opposite interaction effects with host genotype in the *ABO* gene (**Fig 3B-C**).

**Figure 3.**
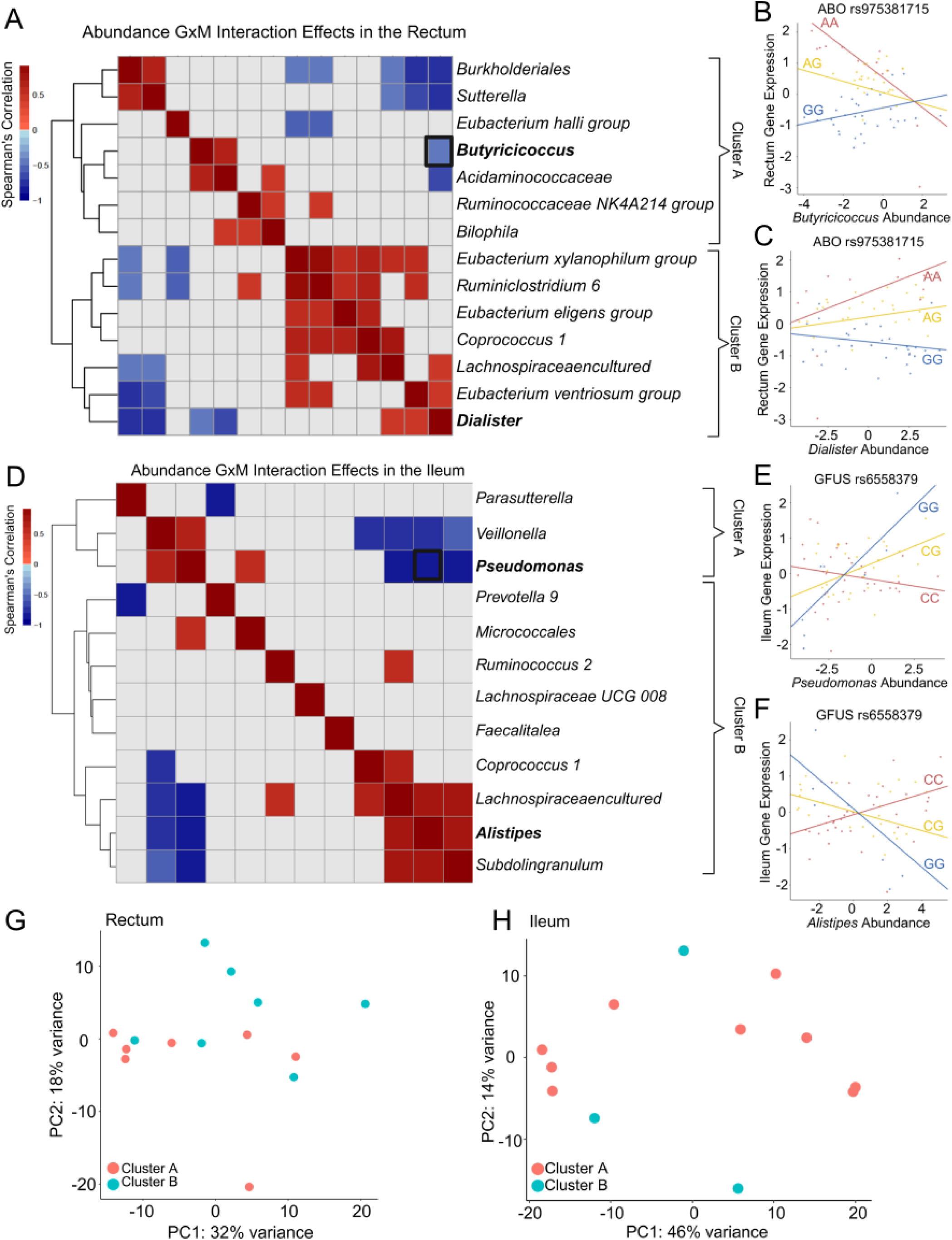
Interactions between host genotype and microbial abundance. **A)** and **D)** Heatmaps of the Spearman correlation of the normalized interaction effects across all host genes that had at least one significant GxM (FDR = 10%) for the microbial taxa with GxM in the rectum **(A)** or in the ileum **(D)**. Nonsignificant correlations are grayed out. Taxa are clustered via hierarchical clustering and cluster into two main groups with cluster A containing microbes that tend to be markers of dysbiosis and cluster B containing more known beneficial microbes in either body site. **B)** Example of GxM for microbial abundance for *Butyricicoccus* in *ABO* gene (FDR = 10%) and **C)** Nominally significant (p < 0.05) example of GxM for abundance of *Dialister* with *ABO* gene in the rectum. Gene Expression residuals are plotted on Y axis and CLR-transformed microbial abundance on the X axis. The taxa and their correlation are bolded in the heatmap **(A)**. **E)** Example of GxM for microbial abundance for *Pseudomonas* in *GFUS* gene (FDR = 10%) and **F)** example of GxM for abundance of *Alistipes* with *GFUS* gene in the ileum (FDR = 10%). Gene Expression residuals are plotted on Y axis and CLR-transformed microbial abundance on the X axis. The taxa are bolded in the heatmap **(D)**. **G)** and **H)** Scatterplots of principal components (PCs) 1 and 2 calculated on CLR-transformed abundance of taxa with GxM in the rectum **(A)** and Ileum **(D)**. The taxa are colored by which cluster they are in in the heatmaps in **A)** and **D)**.

In the ileum, there were also two clusters of taxa with opposite interaction effects. Ileum-cluster A contained three taxa that are known to interact with the host immune system and either promote commensal colonization (*Parasutterella[70])* or are opportunistic pathogens known to promote intestinal inflammation (*Veillonella[71–73]* and *Pseudomonas[74,75]*). Ileum-cluster B however contained more taxa with a variety of metabolic functions in the host which are known to modulate the mucosal environment either through SCFA production[76–79], bile acid utilization[25] and mucin degradation[80] and may be found reduced or elevated in IBD depending on the taxa but appear to primarily interface with the host through metabolites. *Pseudomonas* in Ileum-cluster A and *Alistipes* in Ileum-cluster B had opposing interaction effects with host genotype in the host gene *GFUS*, which encodes a synthase that produces GDP-L-fucose, an essential substrate for fucosylation which is utilized by known IBD risk gene *FUT2* to protect intestinal epithelial barrier function[81].

Previous studies have identified co-abundance networks specific to different diseases including IBD which could be considered as proxies for identifying possible microbial interactions through association with pathway analyses[82]. We therefore investigated whether these clusters of GxM effects on host gene expression reflect clusters of microbial abundances, representative of microbial networks that could be acting in concordance within the gut to modulate host gene regulation. We calculated the principal components of microbial abundance and found that PC1 explained only 32% and 46% of the variance in abundance for the microbes with GxM in the rectum and ileum, respectively. However, when visualizing the microbes positions along PC1 and PC2, we did not observe clustering of the microbes corresponding to the GxM interaction clusters identified earlier (**Fig 3G-H**). This result indicates that taxa with similar GxM effects on host gene expression do not cluster by abundance. Therefore, the clustering of interaction effects could be driven by factors such as the functional niche of each taxa and its role in interfacing with the host, either through metabolites or immune system engagement.

### Host genotypes interact with microbial presence to modulate host gene regulation

IBD-associated changes in gut microbiome composition generally tend to include decreased alpha diversity, with some species becoming highly abundant such as *Veillonellaceae* and *Bacteriodes* and some species becoming depleted such as *Lachnospiraceae* and *Eubacterium* spp. [83–85]. To investigate whether these more pronounced changes in microbial abundance result in additional GxM, we considered the dichotomy of the presence or absence of different microbes. We tested for interaction with the presence of a microbe for 45 taxa in the rectum and we found 8 GxM (8 host genes and 6 taxa; FDR = 10%, Fig **4A**); in the ileum, we tested for the interaction with the presence/absence for 65 taxa and found 9 GxM (6 host genes and 8 taxa, FDR = 10% **Fig 4B**). We found that one of the GxM interactions with microbial abundance of the taxa *Coprococcus 1* in the host gene *PCDHA6* was also discovered in the presence/absence analysis in the ileum (FDR = 10%). Additionally, we found that 7 of the abundance GxM were nominally significant when we tested for the presence of the taxa in the rectum and 9 of the abundance GxM in the ileum were replicated when testing for the presence of the taxa in the ileum (p < 0.05). In both body sites a larger proportion of presence/absence GxM replicated as abundance GxM at nominal p<0.05 (87.5% versus 36.8% in the rectum and 77.7% versus 47.4% in the ileum (**Fig 4A-B**).

**Figure 4.**
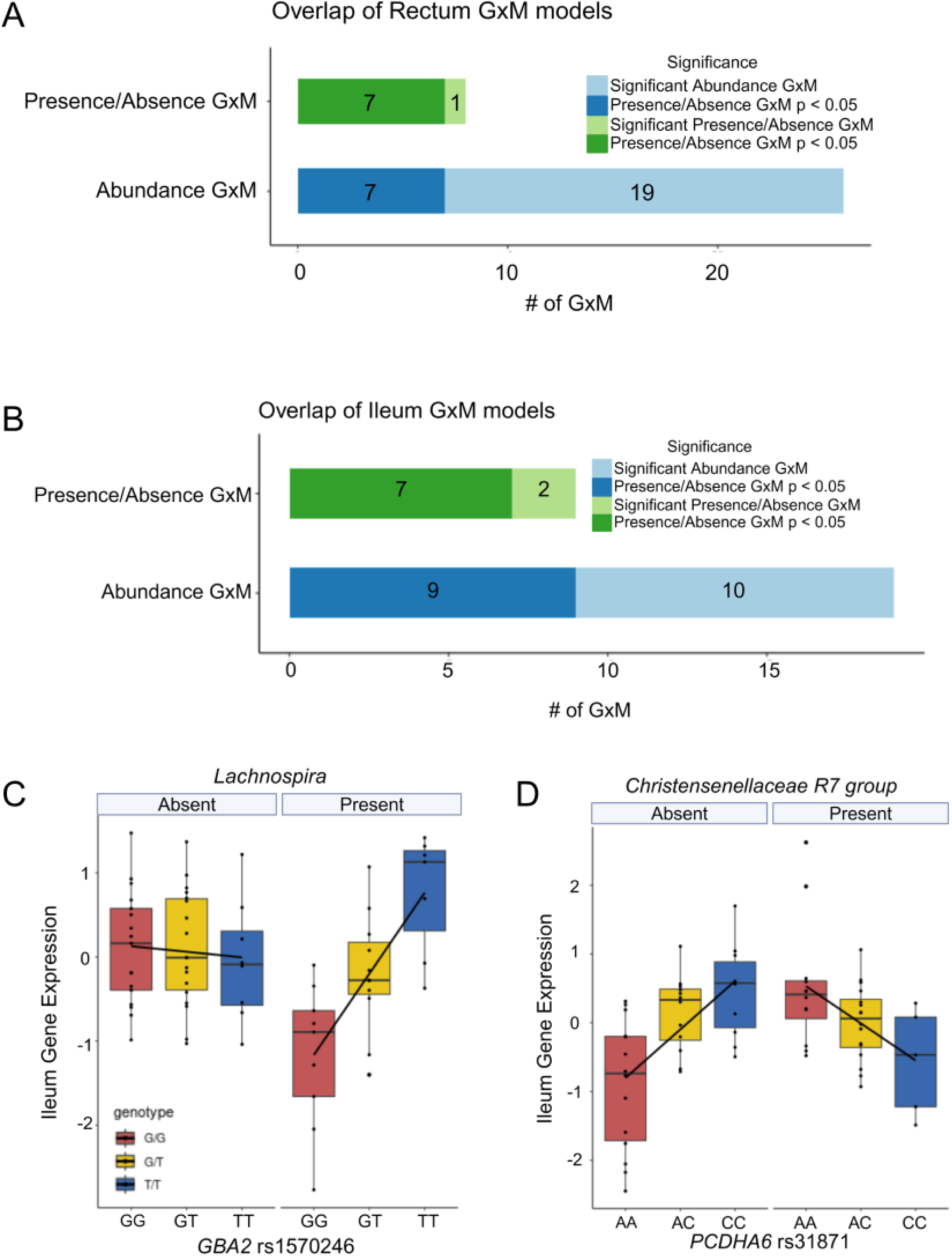
Interactions between host genotype and microbe presence/absence and overlap with abundance GxM. **A-B)** Barplots showing number of GxM identified in the rectum or ileum for each model (abundance and presence/absence) (FDR = 10%) and the overlap with the other model at p-value significance of < 0.05. **C-D)** Examples of GxM for presence/absence of a microbe in the rectum and ileum. Boxplots for each genotype are plotted separated by if the sample had a microbe present or absent with host gene expression residuals plotted on the Y axis. Then within each abundance bin, the black line indicates the line of best fit based on the mean in each genotype group.

Among the presence/absence GxM identified in the rectum, we found that the presence of *Lachnospira* interacted with host genotype to regulate expression of *GBA2*. *GBA2* is a host gene involved in sphingolipid metabolism that was found to be enriched in coding driver mutations in IBD-associated colorectal cancer[86]. In our study we found increased expression of *GBA2* in individuals homozygous for the alternate allele in the presence of the *Lachnospira* (**Fig 4C**). In the ileum, we observed an interaction between the presence of *Christensenellaceae R7 group* and host genotype in *PCDHA6*. *PCDHA6 is* a host gene that encodes a protocadherin, and belongs to a family of proteins important in cell-cell contact and microvilli formation in the small intestine[87].

### Integration of GxM with IBD GWAS data suggest that GxM can modulate IBD risk

As both host genetic variation and the gut microbiome are known risk factors for IBD[4,7,9–11,14,25,28,83–85,88–91], we inquired whether IBD risk can be mitigated or enhanced through interactions between host genotype and microbiome. To identify the contribution of GxM to IBD risk, we performed colocalization of the fine-mapped rectum and ileum eQTLs with the IBD, CD and UC GWAS data from the UK Biobank[88,92], and used SMR[56] to perform transcriptome-wide association study (TWAS). We then used INTACT[57] to integrate TWAS and colocalization results and identify putatively causal host genes. We identified 154 and 131 putatively causal host genes for IBD risk in the rectum and ileum respectively, with 80 genes shared between the two body sites (**Fig 5A**, FDR = 10%).

**Figure 5.**
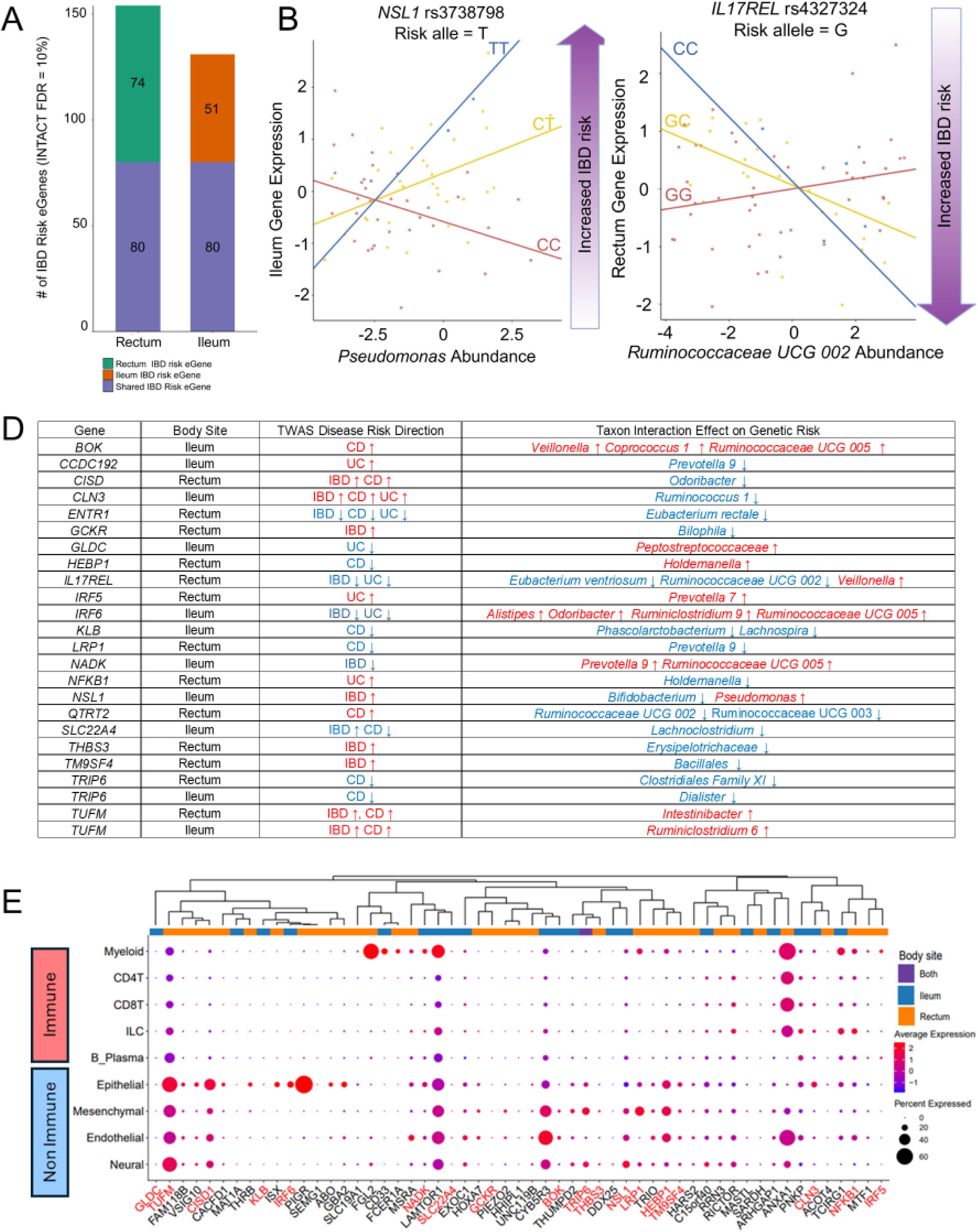
The role of GxM in IBD and different gut cell types. **A)** Barplot showing the number of eGenes identified in the Ileum and Rectum that are associated with IBD risk through INTACT (FDR = 10%) and the overlap between the two body sites. **B-C)** Example of abundance GxM in IBD risk genes in the rectum and ileum. Gene Expression residuals are plotted on Y axis and CLR-transformed microbial abundance on the X axis. The purple arrow indicates the direction of gene expression that is associated with increased IBD risk based on the z-score calculated through INTACT. **D)** Summary of IBD risk genes with GxM: Column 1 is gene; column 2 is body site where genetically predicted gene expression is found to have significant association with disease (TWAS); column 3 indicates effect of genetically predicted gene expression on IBD risk based on TWAS. Text is red if risk is predicted to increase with high expression and is blue if risk is predicted to decrease with high expression; Column 4 indicates the direction that the effect of the microbe has on risk for individuals with the risk allele. Text is red if increased abundance of taxa is associated with increased genetic risk and text is blue if increased abundance of taxa is associated with decreased genetic risk. **E)** Dotplot showing expression of genes with GxM in the rectum and ileum in the major cell types in the scIBD dataset. Size of dots are percent of cells expressing genes and color represents scaled average expression. Genes are ordered based on hierarchical clustering of the scaled average expression of the genes across each cell type and the annotation bar indicates if the gene exhibits GxM in the rectum, ileum or both. Genes in red text are IBD risk genes with GxM based on INTACT and stratified FDR (FDR = 10%).

We then investigated whether any of the host genes that harbored GxM were associated with IBD risk and we found that two genes with GxM in the ileum were associated with IBD risk. For example, increased expression of *NSL1,* a key component of the kinetochore complex in mitosis, was associated with increased risk of IBD. *NSL1* harbored an interaction eQTL with the abundance of *Pseudomonas* where increased abundance of the taxa was associated with increased expression for individuals that harbor the risk allele (T allele) thus amplifying genetic risk (**Fig 5B**). We did not find any overlap in the genes with GxM and IBD risk genes in the rectum at FDR = 10%. As this could be due to a lack of power and the heterogeneity in the data used for this analysis, we then used a stratified FDR approach[58] to identify possible GxM associated with IBD risk genes in both body sites. With this approach we found that 13 of the IBD risk genes in the rectum harbor GxM with 14 taxa for 16 interactions total. In the rectum, we found that expression of *IL17REL*, a gene that enables the IL-17 pathway with known risk variants for Crohn’s Disease[93], was modulated by a GxM between the SNP rs4327324 and the abundance of *Ruminococcaceae UCG 002,* a short chain fatty acid producing microbe found to be reduced in individuals with IBD[94] (**Fig 5C**). Low expression of *IL17REL* is predicted to be associated with increased IBD and UC risk based on TWAS. Our results show that increased abundance of *Ruminococcaceae UCG 002* is associated with increased expression of the gene for individuals homozygous for the risk allele (G allele), thus potentially buffering genetic risk. However, for individuals with the protective allele, high abundance of this taxon was associated with decreased expression, thus diminishing its otherwise protective effect at a lower abundance. In the ileum we found 19 GxM for 11 IBD risk genes and 16 taxa (FDR = 10%). We also found two IBD risk genes, *TRIP6* and *TUFM*, that harbored GxM in both body sites (**Fig 5D**).

To investigate the cellular context of these interactions, we explored the expression of host genes with GxM in different gut cell types from the single cell IBD (scIBD) dataset, which consists of twelve meta-analyzed datasets[59]. We characterized groups of host genes whose expression varies between different major cell compartments: epithelial, endothelial, neural, immune, and mesenchymal cells (**Fig 5E**). We found that 19 host genes with GxM are more highly expressed or detected in a larger percentage of cells in the non-immune lineages versus the immune lineages, particularly the IBD risk genes with GxM (11 host genes). This suggests that interactions between host genetic variation and microbiome in non-immune cell types (e.g. epithelial and mesenchymal) could also contribute to IBD risk.

## Discussion

In this study, we characterized host genetic effects on host gene regulation and their interactions with microbes in the rectum and ileum using data from the IBDMDB cohort and elucidated how these interactions may play a role in IBD risk. We found that eQTLs effects are correlated in rectal and ileum tissue. IBDMDB eQTLs are also enriched for eQTLs in the terminal ileum, transverse and sigmoid colon in the GTEx project, which suggests a common regulatory architecture across these colonic regions. Genetic effects on gene regulation can be tissue and context specific[61,95]. Larger cohorts have been used for eQTL discovery in other colonic sites such as the transverse colon and ileum[61] but studies in the rectum are still limited, despite the rectum being the site for severe disease activity in both CD and UC and is also relevant for other digestive diseases such as colorectal cancer[96]. Furthermore, while large eQTL studies such as GTEx have looked at the ileum, even fewer studies have yet considered the connection of IBD and inflammation when looking at tissue specific effects of gene regulation which does allow for maintaining colonic homeostasis[97]. In our study we identified 288 eGenes that are unique in the IBDMDB ileum dataset and 226 eGenes that are unique in the IBDMDB rectum dataset when compared to GTEx which may indicate tissue-specific or disease specific genetic effects of gene regulation. While larger sample sizes may be necessary to assess disease or tissue specificity, this finding further highlights the necessity to design studies that include not only different tissues, but span multiple contexts such as disease status.

Previous studies have identified host gene regulation that is modified through interactions between host genetic variation and different pathogens including *Mycobacterium tuberculosis* infection, *Listeria monocytogenes*, *Salmonella typhimurium* and Rhinovirus and how these interactions impact host immune response[36,38,39]. Our results demonstrated genetic effects on host gene expression that are modulated by the abundances of commensal microbes. We further showed that different groups of microbes, characterized by being either beneficial or opportunistic pathogens, can have opposing roles in the gut. For example, we found opposing interaction effects between *Butyricicoccus* and *Dialister* at the eQTL rs975381715 for the host gene *ABO,* which determines blood type. Both taxa are short chain fatty acid producers, but *Butyricococcus* specifically produces butyrate while *Dialister* produces propionate through succinate consumption[98,99]. Previously, the *ABO* locus has been associated with interindividual differences in gut microbiome composition[100] potentially through altering N-acetyl-galactosaminyl-transferase levels in a genotype dependent manner[101]. Butyrate has been found to induce expression of glycosyltransferase genes such as N-acetyl-galactosaminyl-transferase supporting the notion that SCFAs may modulate glycan biosynthesis[102]. Thus, the opposing interaction effects observed may reflect differences in how butyrate and propionate influence mucosal glycosylation thereby modulating the effect of *ABO* genotype on host gene expression. Furthermore, these specific microbes have not been linked to the *ABO* gene in previous studies, highlighting that integrating host gene regulation may reveal host genotype-microbe interactions that may be missed when solely looking at microbial abundance and host genetic variation.

Prior studies have elucidated how the gut microbiome interacts with different cell types including immune cells[103] and epithelial cells[104] and how these reciprocal interactions maintain intestinal homeostasis. By integrating our results with the scIBD dataset,[59] we found that genes with GxM tend to be more highly expressed in non-immune lineages, particularly epithelial cells. IBD pathology is classically attributed to hyper-reactivity of both the innate and adaptive immune system[105,106]. However, studies have shown that IBD is also associated with altered physiological processes in non-immune gut cell types, including breakdown of the epithelium barrier and tissue remodeling by stromal cells[107,108]. Prior single cell RNAseq studies showed that epithelial and stromal cells acquire a pro-inflammatory signature that could contribute to IBD pathology. Additionally, monogenic causes of IBD primarily involve disruption of epithelial barrier integrity-related processes[109,110]. In our study we found that IBD risk genes with GxM tend to have higher expression in non-immune or non-hematopoietic cell lineages. This suggests that GxM is a plausible mechanism underlying the development of initial epithelial/mesenchymal inflammation or injury previously reported in several studies of IBD initiation[111]. The standard of care for IBD usually involves use of biologic treatments aimed at reducing inflammation through targeting immune effector cells to allow for the ability for the intestinal epithelium to heal[112]. Our findings indicate that jointly considering host genetic variation and microbiome composition could allow for the discovery of other therapeutic targets especially in the epithelium and mesenchyme.

Our statistical power to detect tissue specific eQTLs and GxM is limited by the number of individuals with paired genotype, transcriptomic and microbial 16S data which was a smaller subset of the larger cohort. We only found 310 SNP-gene pairs that harbor eQTLs in both body sites, and this is likely due to the small sample size and limited statistical power. While we tested for both the abundance and presence/absence of a microbe in the respective tissue sites we find a larger proportion of presence absence GxM tends to replicate in the abundance model at the nominal p-value level. This indicates that the abundance GxM model can potentially also pick up these dichotomous interactions if afforded enough statistical power and this could be remedied through larger cohorts designed to investigate the role of GxM in host gene regulation. Another limitation is that for the microbial data we only have 16S rRNA data which is low resolution and thus limits the specificity of the host genotype-taxa relationships we were able to identify, but this limitation should not result in GxM false positives. By integrating host transcriptomic data with metagenomic data collected from tissues we could identify GxM at the microbial strain level. This would provide testable microbial targets that would be invaluable in our understanding of how different species/strains contribute to physiology as well as to disease pathology. In addition, while we were able to elucidate if any of the genes with GxM were highly expressed in different cell types, there is still a need for studies where microbial composition and single-cell resolution data are collected in order to identify interactions between host genotype, cell type and microbial composition (GxCxM).

The identification of rectum and ileum eQTLs in host genes known to harbor risk variants associated with IBD through TWAS and colocalization with GWAS further highlights the link between genetic regulation of gene expression and susceptibility to IBD. Through integration of GxM with GWAS and TWAS data we can characterize their clinical significance. Fecal microbiota transplants (FMTs) have been proposed as a potential therapeutic for IBD[113]. While this approach has been effective for treating *Clostridium difficile* infection, its success in IBD has been variable and appears to depend on the donor and preparation of the FMT[114]. Using specific strains with well characterized host genotype-specific effects could potentially improve therapeutic outcomes in IBD[115]. For example, we found that *IL17REL* harbors a GxM at rs4327324 (G>C) where individuals with the protective allele (C), and increased abundance of *Ruminoccocaceae UCG 002,* an SCFA producing microbe, exhibit decreased *IL17REL* expression, in a direction consistent with increased disease risk. Thus, even microbes considered beneficial may have different effects depending on host genotype highlighting the need for personalized microbial therapeutics that consider interactions between host genetic variation and microbial composition.

## Conclusions

In summary, these findings provide evidence of how host genetic variation and the gut microbiome can jointly regulate host gene expression in the rectum and ileum which has translational implications for understanding and treating IBD. The connections between genetic variation, microbiome, and disease phenotype offer a multidimensional perspective that could shape future research directions and clinical applications in the field of gastroenterology and precision medicine.

## Data Availability

The data used for the analyses described in this manuscript were obtained from the
Inflammatory Bowel Disease Multi'Omics Database website and the Human Microbiome Project via dbGaP accession phs001626.v1.p1. Data from the GTEx Consortium was also used for the analysis in this manuscript and was obtained from the GTEx Portal and the GTEx Consortium via dbGaP accession phs000424.v9.p2.
Summary statistics for the data analysis are available as supplemental materials in the manuscript.

https://ibdmdb.org

## List of Abbreviations

IBD: Inflammatory Bowel Disease
GxM: Genotype x Microbiome interaction
eQTL: expression Quantitative Trait Loci
CD: Crohn’s Disease
UC: Ulcerative Colitis
GWAS: Genome-Wide Association Studies
TWAS: Transcriptome-Wide Association Studies
FDR: False Discovery Rate
SNP: Single Nucleotide Polymorphism
GxE: Genotype x Environment Interaction
GTEx: Gene-Tissue Expression Consortium
OR: Odds Ratio
SCFA: Short Chain Fatty Acid
IBDMDB: Inflammatory Bowel Disease Multi’Omics Database
scIBD: Single Cell Inflammatory Bowel Disease dataset
FMT: Fecal Microbiota Transplant
GxCxM: Genotype x Cell Type x Microbiome interaction

## Availability of Data and Materials

The data used for the analyses described in this manuscript were obtained from the Inflammatory Bowel Disease Multi’Omics Database website (https://ibdmdb.org) and the Human Microbiome Project via dbGaP accession phs001626.v1.p1. Data from the GTEx Consortium was also used for the analysis in this manuscript and was obtained from the GTEx Portal (https://gtexportal.org) and the GTEx Consortium via dbGaP accession phs000424.v9.p2. Summary statistics for the data analysis are available as supplemental materials. The code and scripts used in data analysis are available on GitHub (https://github.com/piquelab/GxM_IBD).

## Ethics declarations

This study was determined to qualify for Exemption according to category 4 by the Wayne State University IRB (Protocol # IRB-22-05-4638).

## Consent for publication

Not applicable

## Author Information

### Contributions

SN downloaded the data, analyzed the data, interpreted the results, prepared the figures, wrote the first draft of the manuscript and revised the manuscript. SA provided data analysis scripts, analyzed the data, interpreted the results, provided feedback on the first draft of the manuscript. SP provided data analysis scripts and provided feedback on the first draft of the manuscript. JW analyzed the data, wrote parts of the manuscript and provided feedback on the first draft of the manuscript. RB supervised the project, interpreted the results, provided feedback on the manuscript, revised the manuscript. RPR supervised the project, interpreted the results, provided feedback on the manuscript, revised the manuscript. FL designed the project, provided funding, supervised the project, interpreted the results, provided feedback on the manuscript, revised the manuscript. All authors have read and approved the final manuscript.

## Acknowledgements

We thank members of the Luca/Pique-Regi and Blekhman groups, and Xiaoquan Wen for helpful comments and discussions. The IBDMDB was conducted by the IBDMDB Investigators and supported by the National Institute of Diabetes and Digestive and Kidney Diseases (NIDDK), National Human Genome Research Institute (NHGRI), National Center for Advancing Translational Sciences (NCATS), National Science Foundation (NSF), Army Research Office, Swedish Research Council, Helen Hay Whitney Foundation, Crohn’s and Colitis Foundation, and Helmsley Charitable Trust. The data from the IBDMDB reported here were supplied by investigators at the Broad Institute, Massachusetts General Hospital, Cincinnati Children’s Hospital, Emory University Hospital, and Cedars-Sinai Medical Center. This manuscript was not prepared in collaboration with the IBDMDB study investigators and does not necessarily reflect the opinions or views of the IBDMDB study, the NIDDK, NHGRI, NCATS, NSF, Army Research Office, Swedish Research Council, Helen Hay Whitney Foundation, Crohn’s and Colitis Foundation, or Helmsley Charitable Trust. Figures in this manuscript were created and/or formatted using <u>BioRender.com</u>.

## Supplemental Figures

**Supplemental Fig 1.**
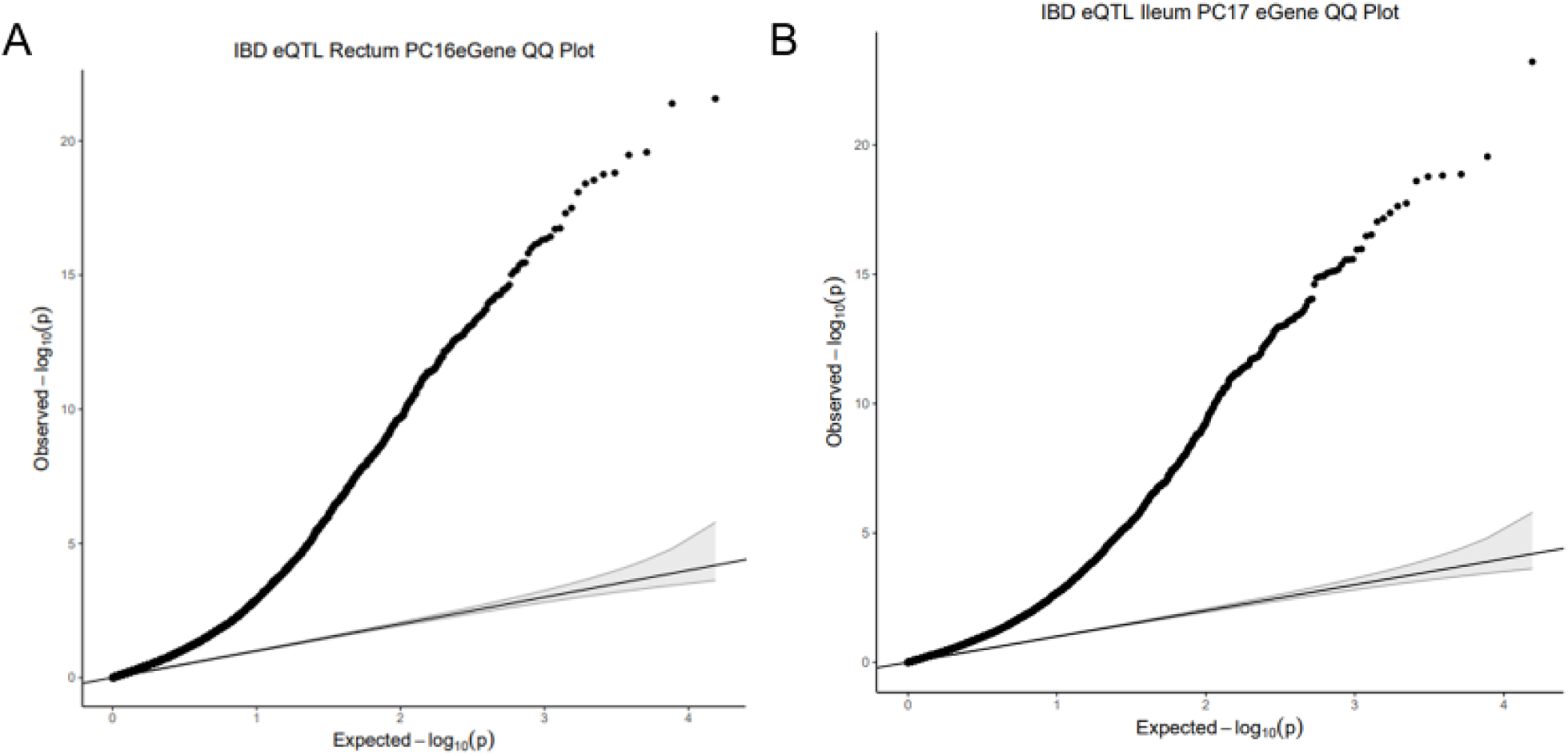
QQ (quantile-quantile) plots of P values from FastQTL for the A) Rectum eQTL analysis after correcting for 16 gene expression PCs and B) the Ileum eQTL analysis after correcting for 17 gene expression PCs.

**Supplemental Figure 2.**
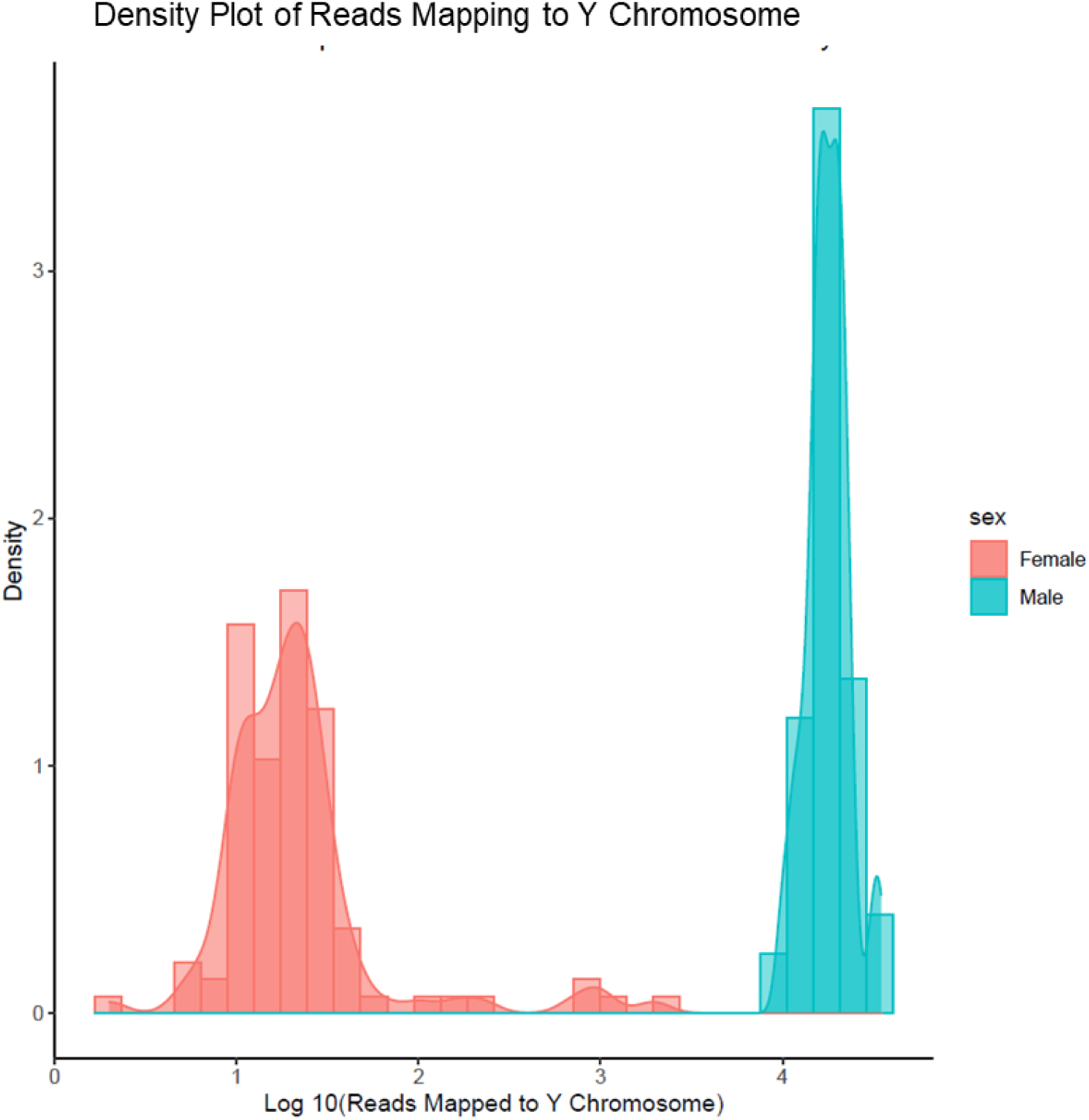
Density plot of reads in RNA seq data mapping to Y chromosome colored by self-reported sex

**Supplemental Figure 3.**
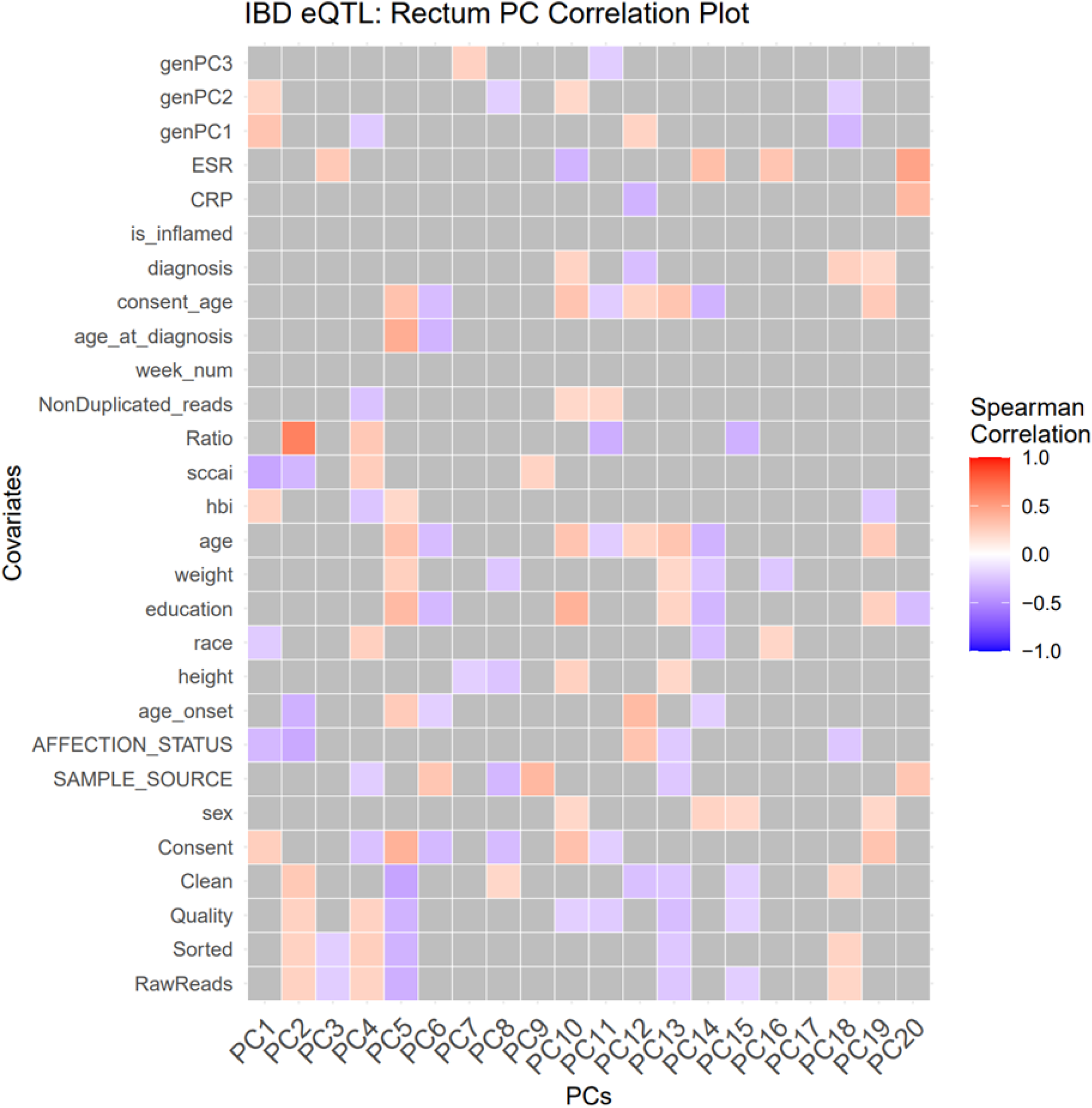
Heatmap of the Spearman correlation of the first 20 PCs of gene expression data from the rectum with different covariates including technical and cohort characteristics. Nonsignificant correlations are grayed out

**Supplemental Figure 4.**
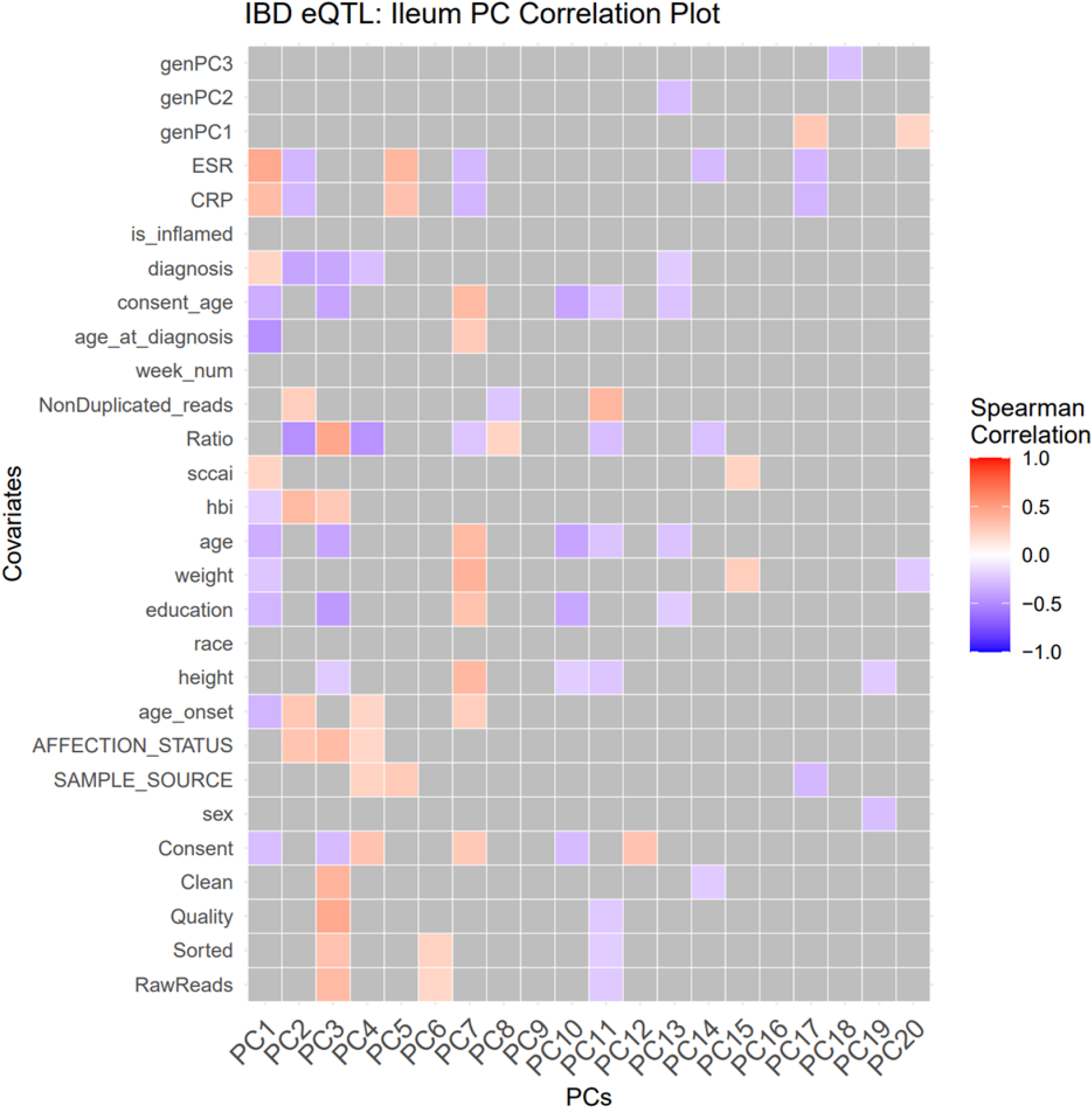
Heatmap of the Spearman correlation of the first 20 PCs of gene expression data from the ileum with different covariates including technical and cohort characteristics. Nonsignificant correlations are grayed out

**Supplemental Figure 5.**
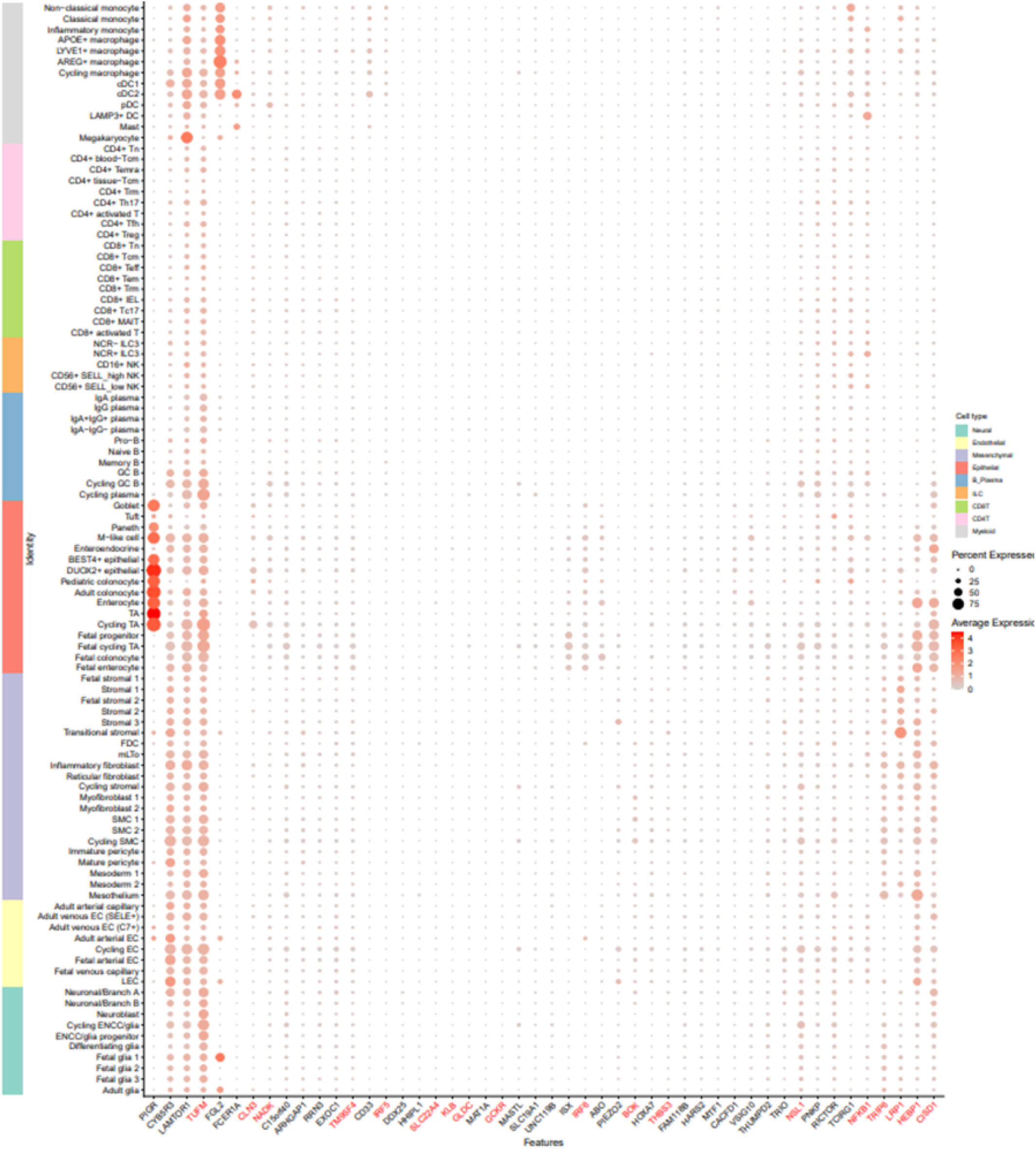
Dotplot showing expression of genes with GxM in the rectum and ileum in the scIBD dataset. Genes in red text are IBD risk genes with GxM based on INTACT and stratified FDR (FDR = 10%). Size of dots are percent of cells expressing genes and color represents normalized average expression. Dendrogram at the top of the figure is generated using pheatmap based on hierarchical clustering of the average expression of the genes across each cell type and the annotation bar indicates if the gene exhibits GxM in the rectum or ileum. Colored bar to left of plot indicates major cluster each individual cell type belongs to.

## Supplemental Tables

**Supplemental Table 1 Metadata for Ileum and Rectum Samples.** This table contains the metadata for all the RNA-seq samples used in this study. Columns: IBDMBD Participant ID, Project ID, Body Site, Diagnosis

**Supplemental Table 2 Rectum eQTL Summary Statistics**. Results of FastQTL with adaptive permutations after correcting for 16 Gene Expression PCs. Columns: pid - ensemble gene ID, sid - lead SNP rsID, dist - distance between SNP and TSS, npval - the nominal p-value (p-value before adaptive permutations), slope - effect size of variant on gene expression, ppval - permutation-corrected p-value, bpval - Beta-approximated empirical p-value, bqval - Final FDR calculated using Storey’s q-value method on the Beta-approximated p-value

**Supplemental Table 3 Fine-mapped SNP-Gene Pairs in Rectum.** SNP with the top PIP from DAP-G for each eGene identified through FastQTL in the rectum. Columns: pid - ensemble gene ID, sid - rsID for SNP with top PIP.

**Supplemental Table 4 Ileum eQTL Summary Statistics**. Results of FastQTL with adaptive permutations after correcting for 17 Gene Expression PCs. Columns: pid - ensemble gene ID, sid - lead SNP rsID, dist - distance between SNP and TSS, npval - the nominal p-value (p-value before adaptive permutations), slope - effect size of variant on gene expression, ppval - permutation-corrected p-value, bpval - Beta-approximated empirical p-value, bqval - Final FDR calculated using Storey’s q-value method on the Beta-approximated p-value.

**Supplemental Table 5 Fine-mapped SNP-Gene Pairs in Ileum**. SNP with the top PIP from DAP-G for each eGene identified through FastQTL in the ileum. Columns: pid - ensemble gene ID, sid - rsID for SNP with top PIP.

**Supplemental Table 6 GxM Results for Presence/Absence of a Microbe in Rectum**. Results from the linear model testing for interaction between genotype and presence/absence of a microbe for all microbes and all SNP-gene pairs tested in the Rectum. Columns: pid - ensemble gene ID, sid - rsID for SNP, Intercept_pval - nominal p-value for the intercept, dosage_pval - nominal p-value for the genotype effect, microbe_pval - nominal p-value for microbe effect, interaction_pval - nominal p-value for the interaction term, Intercept_effect - estimated regression coefficient for the intercept, dosage_effect - estimated regression coefficient for the genotype, microbe_effect - estimated regression coefficient for microbe effect, interaction_effect - estimated regression coefficient for the interaction term, Intercept_SE - standard error for intercept effect, dosage_SE - standard error for genotype effect, microbe_SE - standard error for microbe effect, interaction_SE - standard error for interaction term, Intercept_qval - Storey’s q-value for intercept, dosage_qval - Storey’s q-value for genotype effect, microbe_qval - Storey’s q-value for microbe effect, interaction_qval - Storey’s q-value f or the interaction term, microbe - taxa tested for the interaction, symbol - gene symbol.

**Supplemental Table 7 GxM Results for Presence/Absence of a Microbe in Ileum**. Results from the linear model testing for interaction between genotype and presence/absence of a microbe for all microbes and all SNP-gene pairs tested in Ileum. Columns: pid - ensemble gene ID, sid - rsID for SNP, Intercept_pval - nominal p-value for the intercept, dosage_pval - nominal p-value for the genotype effect, microbe_pval - nominal p-value for microbe effect, interaction_pval - nominal p-value for the interaction term, Intercept_effect - estimated regression coefficient for the intercept, dosage_effect - estimated regression coefficient for the genotype, microbe_effect - estimated regression coefficient for microbe effect, interaction_effect - estimated regression coefficient for the interaction term, Intercept_SE - standard error for intercept effect, dosage_SE - standard error for genotype effect, microbe_SE - standard error for microbe effect, interaction_SE - standard error for interaction term, Intercept_qval - Storey’s q-value for intercept, dosage_qval - Storey’s q-value for genotype effect, microbe_qval - Storey’s q-value for microbe effect, interaction_qval - Storey’s q-value f or the interaction term, microbe - taxa tested for the interaction, symbol - gene symbol.

**Supplemental Table 8 GxM Results for Abundance of a Microbe in Rectum**. Results of the linear model testing for interaction between genotype and microbial abundance for all microbes and all SNP-gene pairs tested in the Rectum and Ileum. Columns: pid - ensemble gene ID, sid - rsID for SNP, Intercept_pval - nominal p-value for the intercept, dosage_pval - nominal p-value for the genotype effect, microbe_pval - nominal p-value for microbe effect, interaction_pval - nominal p-value for the interaction term, Intercept_effect - estimated regression coefficient for the intercept, dosage_effect - estimated regression coefficient for the genotype, microbe_effect - estimated regression coefficient for microbe effect, interaction_effect - estimated regression coefficient for the interaction term, Intercept_SE - standard error for intercept effect, dosage_SE standard error for genotype effect, microbe_SE - standard error for microbe effect, interaction_SE - standard error for interaction term, Intercept_qval - Storey’s q-value for intercept, dosage_qval - Storey’s q-value for genotype effect, microbe_qval - Storey’s q-value for microbe effect, interaction_qval - Storey’s q-value f or the interaction term, microbe - taxa tested for the interaction, symbol - gene symbol.

**Supplemental Table 9 GxM Results for Abundance of a Microbe in Ileum**. Results of the linear model testing for interaction between genotype and microbial abundance for all microbes and all SNP-gene pairs tested in the Ileum. Columns: pid - ensemble gene ID, sid - rsID for SNP, Intercept_pval - nominal p-value for the intercept, dosage_pval - nominal p-value for the genotype effect, microbe_pval - nominal p-value for microbe effect, interaction_pval - nominal p-value for the interaction term, Intercept_effect - estimated regression coefficient for the intercept, dosage_effect - estimated regression coefficient for the genotype, microbe_effect - estimated regression coefficient for microbe effect, interaction_effect - estimated regression coefficient for the interaction term, Intercept_SE - standard error for intercept effect, dosage_SE standard error for genotype effect, microbe_SE - standard error for microbe effect, interaction_SE - standard error for interaction term, Intercept_qval - Storey’s q-value for intercept, dosage_qval - Storey’s q-value for genotype effect, microbe_qval - Storey’s q-value for microbe effect, interaction_qval - Storey’s q-value f or the interaction term, microbe - taxa tested for the interaction, symbol - gene symbol.

**Supplemental Table 10 INTACT IBD Risk Genes in the Rectum**. Results of INTACT analysis to identify IBD risk genes in Rectum. Columns: ensgene - ensemble gene ID, GRCP - Gene-level region colocalization probability, GLCP - Gene-level Locus Colocalization Probability, zscore - GWAS z-score, LFDR - Local False Discovery Rate, FDR - Benjamini-Hochberg False Discovery Rate across all genes, Trait - UK Biobank IBD GWAS trait, symbol - gene symbol.

**Supplemental Table 11 INTACT IBD Risk Genes in the Ileum**. Results of INTACT analysis to identify IBD risk genes in Rectum and Ileum. Columns: ensgene - ensemble gene ID, GRCP - Gene-level region colocalization probability, GLCP - Gene-level Locus Colocalization Probability, zscore - GWAS z-score, trait - UK Biobank IBD GWAS trait which gene is identified to be associated with through INTACT, LFDR - Local False Discovery Rate, FDR – Benjamini-Hochberg False Discovery Rate across all genes.

**Supplemental Table 12 IBD Risk Gene GxM Results for Rectum**. IBD risk genes with significant GxM after using stratified FDR approach for the Rectum. Columns: sid - rsID for SNP, pid - ensemble gene ID, symbol - gene symbol, Trait - UK Biobank IBD GWAS trait, slope - effect size of genotype on gene expression from FastQTL, Intercept_pval - nominal p-value for the intercept, dosage_pval - nominal p-value for the genotype effect, microbe_pval - nominal p-value for microbe effect, interaction_pval - nominal p-value for the interaction term, Intercept_effect - estimated regression coefficient for the intercept, dosage_effect - estimated regression coefficient for the genotype, microbe_effect - estimated regression coefficient for microbe effect, interaction_effect - estimated regression coefficient for the interaction term, Intercept_SE - standard error for intercept effect, dosage_SE - standard error for genotype effect, microbe_SE - standard error for microbe effect, interaction_SE - standard error for interaction term, Intercept_qval - Storey’s q-value for intercept, dosage_qval - Storey’s q-value for genotype effect, microbe_qval - Storey’s q-value for microbe effect, interaction_qval - Storey’s q-value f or the interaction term, strat_interaction_qval - stratified Benjamini Hochberg FDR, microbe - taxa tested for the interaction, GxM - type of GxM identified

**Supplemental Table 13 IBD Risk Gene GxM Results for Ileum**. IBD risk genes with significant GxM after using stratified FDR approach for the Ileum. Columns: sid - rsID for SNP, pid - ensemble gene ID, symbol - gene symbol, Trait - UK Biobank IBD GWAS trait, slope - effect size of genotype on gene expression from FastQTL, Intercept_pval - nominal p-value for the intercept, dosage_pval - nominal p-value for the genotype effect, microbe_pval - nominal p-value for microbe effect, interaction_pval - nominal p-value for the interaction term, Intercept_effect - estimated regression coefficient for the intercept, dosage_effect - estimated regression coefficient for the genotype, microbe_effect - estimated regression coefficient for microbe effect, interaction_effect - estimated regression coefficient for the interaction term, Intercept_SE - standard error for intercept effect, dosage_SE - standard error for genotype effect, microbe_SE - standard error for microbe effect, interaction_SE - standard error for interaction term, Intercept_qval - Storey’s q-value for intercept, dosage_qval - Storey’s q-value for genotype effect, microbe_qval - Storey’s q-value for microbe effect, interaction_qval - Storey’s q-value f or the interaction term, strat_interaction_qval - stratified Benjamini Hochberg FDR, microbe - taxa tested for the interaction, GxM - type of GxM identified

